# C Reactive Protein based early intervention for home quarantined COVID-19 patients to avoid complications and hospitalization

**DOI:** 10.1101/2021.06.22.21258712

**Authors:** Manimarane Arjunan, U.R Anoop, Kavita Verma, Oviyaa Manimarane

**Author notes:** The author is the principal Investigator who provided medical assessment and treatment (Manimarane Arjunan). All authors contributed equally to the study. **Correspondence to:** Anoop U.R, UR Anoop Research Group, Pondicherry, India. 605008. **Funding:** No funding.

## Abstract

**Background:** As India was reeling under the relentless onslaught of the second wave of COVID -19, the hospitals were over-crowded and resources were inaccessible. Many COVID -19 positive patients had to stay at home and reach out to their family physicians for guidance. This gave us an opportunity to observe a group of patients in home quarantine. The challenge was daunting. We could not afford to allow these patients to deteriorate as hospital beds and oxygen support were not readily available. Therefore, we developed an innovative approach to monitor these patients and to intervene at the optimal time to avoid hospitalization.

Based on objective biochemical tests like elevated CRP and clinical signs like persistent high-grade fever, we initiated early low dose steroids and antibiotics prior to the onset of hypoxia. None of the home quarantined patients deteriorated because of the timely medical intervention. These patients neither required oxygen support nor hospitalization. We therefore present our new treatment model for home quarantined COVID-19 patients along with our suggestions for implementing the model at the community level.

**Methods:** 25 patients under home quarantine who contacted online for medical guidance underwent clinical and biochemical evaluation. CT radiological examination was done whenever indicated. Treatments were initiated under medical guidance based on the CRP values and clinical presentation.

**Results:** Among the 25 patients, 40% of the patients (10/25) had mild symptoms with normal CRP. 24% of the patients (6/25) had mild symptoms with elevated CRP. 36% of patients (9/25) had elevated CRP with persistent high fever. All the 25 patients recovered over the next four weeks.

**Conclusion:** We propose an early objective assessment of inflammatory markers like CRP prior to the onset of hypoxia during home quarantine and early, need-based intervention under medical guidance to avoid complications and hospitalization.

## INTRODUCTION

As India was reeling under the relentless onslaught of the second wave of COVID -19, the healthcare system was stretched to its limits. Even after providing exclusive COVID treatment hospitals and increasing the number of beds in private and government hospitals, the number of cases outnumbered the availability of beds, especially during the peak of the second wave.

To avoid the chaos associated with the admission of all COVID positive patients in hospitals, the governments of all affected countries have published guidelines for home quarantine of asymptomatic and mild cases^1^. But these guidelines are mostly based on subjective symptoms.

Currently, the patients reporting at the COVID centers are classified into mild cases based on their clinical presentation which can be misleading. Unfortunately, a large number of patients who were diagnosed as mild cases and advised to be on home quarantine lost their lives either at home or immediately after they were brought to the emergency department because of the delay or failure to identify the danger signs and symptoms of clinical deterioration on time.

Therefore, the success of home quarantine will depend on early identification of the progress of the disease and initiation of the optimal drugs at the right time.

As logistics and availability of resources were disrupted, a high number of patients were facing hardships in getting admissions at the designated COVID centers. Therefore, many patients staying at home had to seek the help of their family physicians.

In this regard, we had the opportunity to provide online consultation to a few home quarantined patients and follow them up for a period of four weeks. During this period, we assessed their clinical signs and symptoms, biochemical markers, oxygen saturation and thoracic CT scan. We noted that some patients who were probably prone to impending clinical deterioration and death could be identified on the third and fourth day of illness by detecting markedly elevated CRP levels.

Because of the increased mortality due to delay in treatment of home quarantined patients, there was a need for an innovative approach to treat patients without allowing any clinical deterioration. It was a daunting challenge. We could not afford to allow the patients to worsen as the hospital beds and oxygen support were not readily available. We therefore focused on clinical signs of high-grade fever and inflammatory markers, particularly CRP levels on the third and fourth day after the onset of symptoms.

We decided to treat them promptly well before the appearance of classical red flag signs like difficulty in breathing, SpO2 <94% on room air, persistent pain or pressure in chest and mental confusion. Particularly, we did not wait for the oxygen saturation to fall to low levels to initiate the interventions.

## MATERIALS AND METHODS

### Aim

1. To study whether early CRP estimation on the third or fourth day in home quarantined COVID-19 patients can identify the patients susceptible to clinical deterioration.
2. To study whether early medical intervention prior to onset of hypoxia, on the third or fourth day in home quarantined COVID-19 patients with elevated CRP can avoid clinical deterioration.
3. To evaluate the number of patients developing coagulation abnormalities as evidenced by elevated D Dimer.

### Inclusion Criteria

1. COVID-19 positive patients under home quarantine
2. Patients consulting online on the 3^rd^ or 4^th^ day after the onset of symptoms.
3. Patients with oxygen saturation 94% and above.

### Exclusion Criteria

1. Patients consulting online on or after 5^th^ day of onset of symptoms.
2. Patients with oxygen saturation less than 94% during the initial presentation.
3. Systemic disease known to increase CRP levels.

### Primary Outcome Measure

1. Number of home quarantined COVID 19 positive patients requiring admission in hospital for hypoxia

Description: Oxygen saturation in patients was monitored using pulse oximetry at home. Those patients who developed hypoxia (less than 94%) were referred for hospitalization.

Time Frame: Two weeks from the onset of symptoms.

### Secondary Outcome Measure

1. Number of home quarantined COVID 19 positive patients developing thromboembolic complications

Description: Thromboembolic complications in COVID-19 positive patients under home quarantine were assessed using an online questionnaire.

Time Frame: Four weeks from the time of initial presentation.

### Data Collection

During the peak of the second wave of the COVID-19 pandemic, patients under home quarantine who contacted online for medical guidance underwent clinical and biochemical evaluation. CT radiological examination was done whenever indicated. All the patients underwent complete blood count, CRP, D dimer, Ferritin, and LDH on the third or fourth day of illness. For mild cases, CRP and D dimer were repeated after three days. For patients who had marked elevation of the inflammatory markers, CRP and D dimer were repeated every third day till the parameters normalized. CT thorax was done for patients with persistent fever on the third day or persistent cough with expectoration. The clinical, biochemical and radiological data were recorded with the consent of the patients.

### Classification of Patients

The cases were classified into three groups based on clinical signs and elevated CRP, namely:

a. Patients with mild symptoms and normal CRP.
b. Patients with mild symptoms and less than 10-fold increase in CRP.
c. Patients with high grade fever persisting even on the third or fourth day after onset of symptoms or 10-fold or more increase in CRP.

### Statistical Analysis

A group of patients under home quarantine were observed (Fig 2,3,4,5,6) with their consent. The group consisted of 25 patients in the age group of 19 years to 72 years with an average of 52 years. In this group, 12 were males and 13 were females.

**FIGURE:1.**
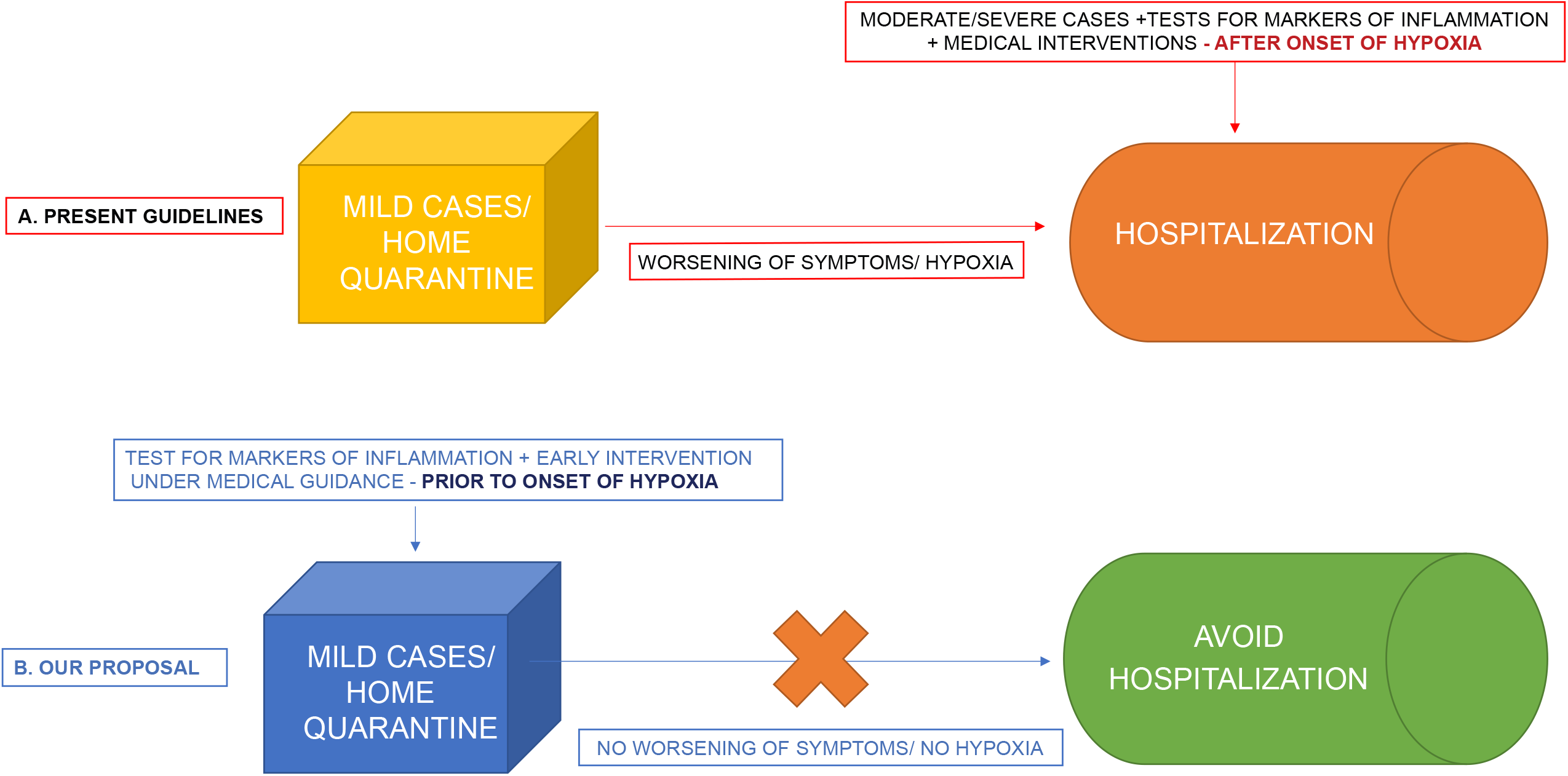

**FIGURE:2.**
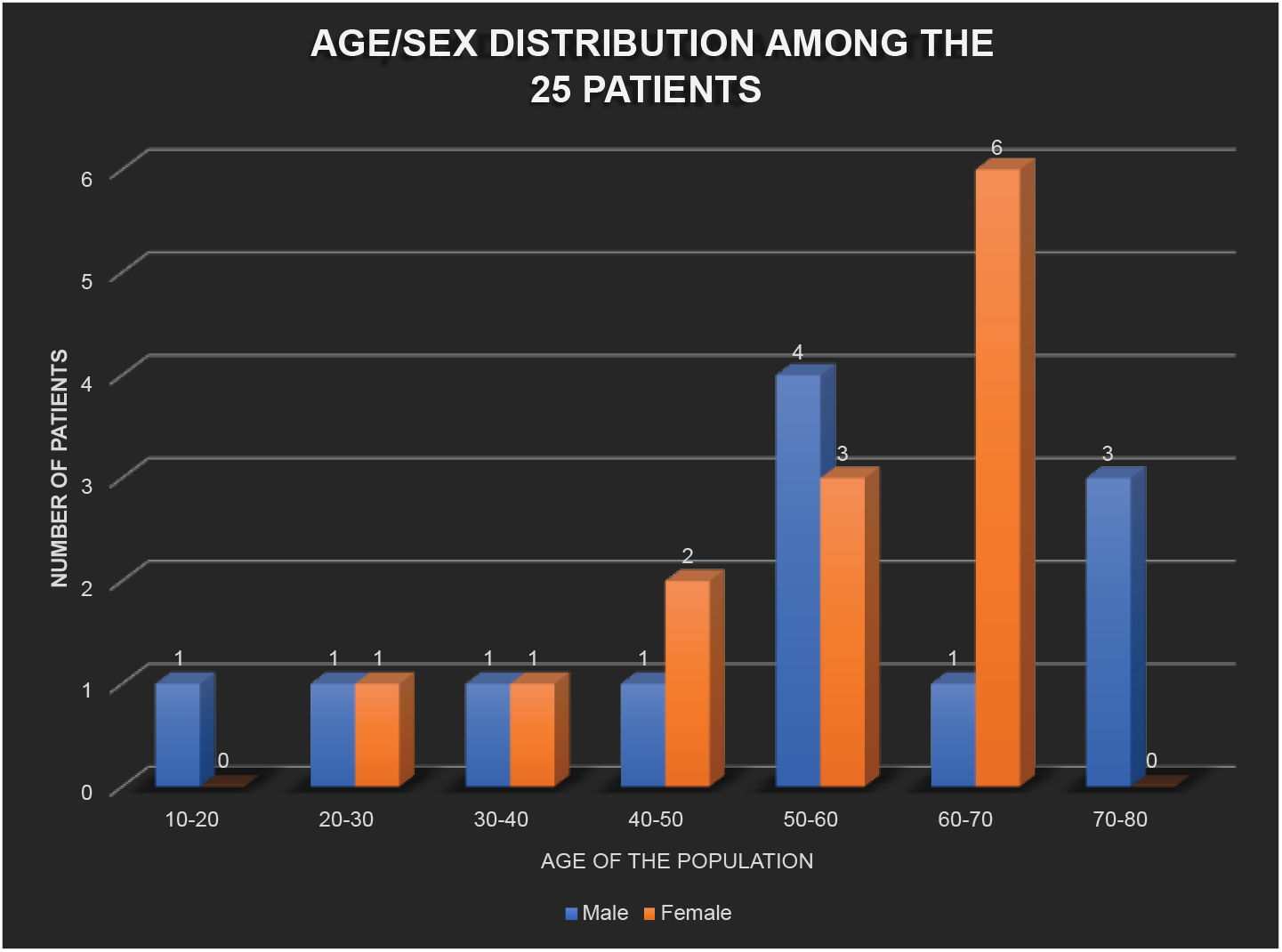

**FIGURE:3.**
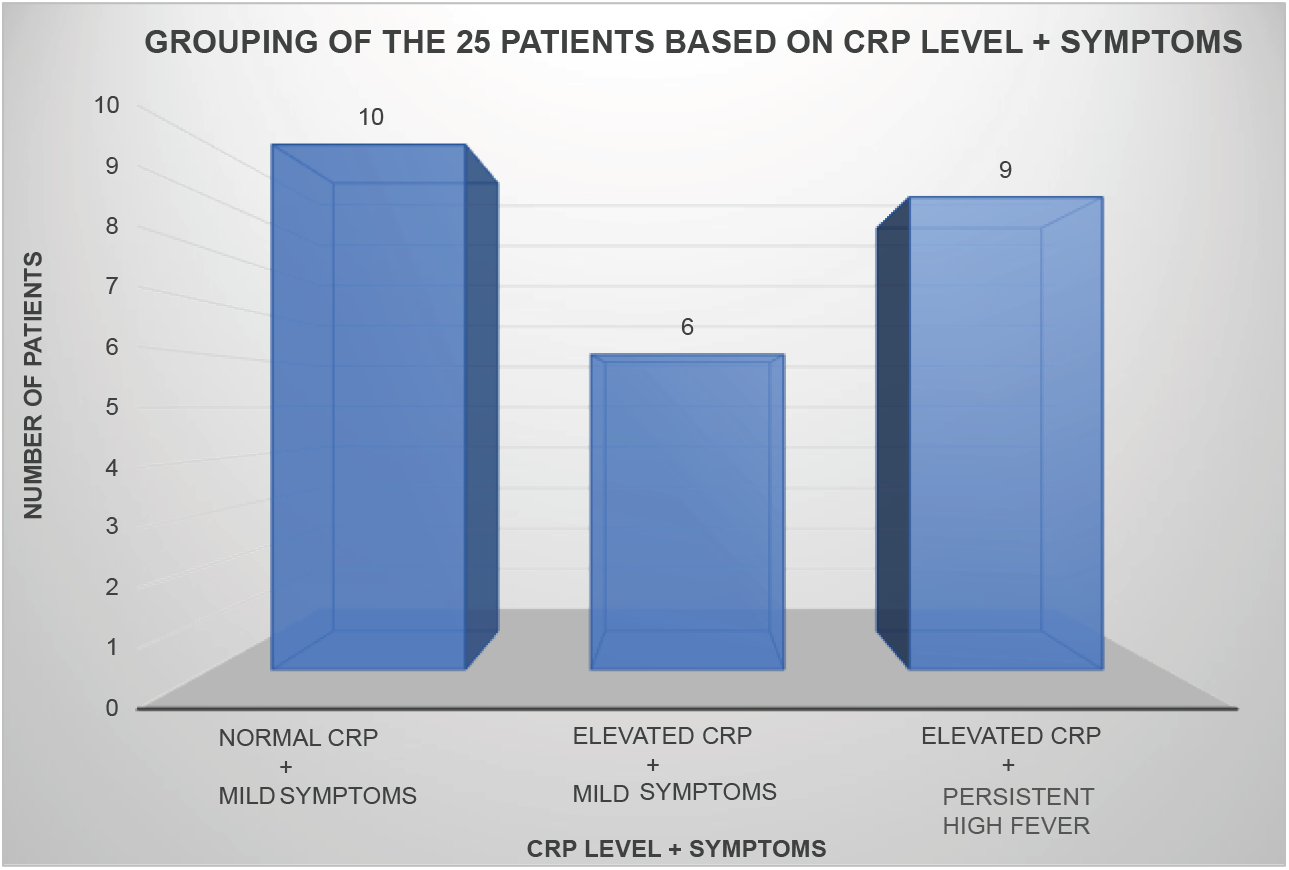

**FIGURE:4.**
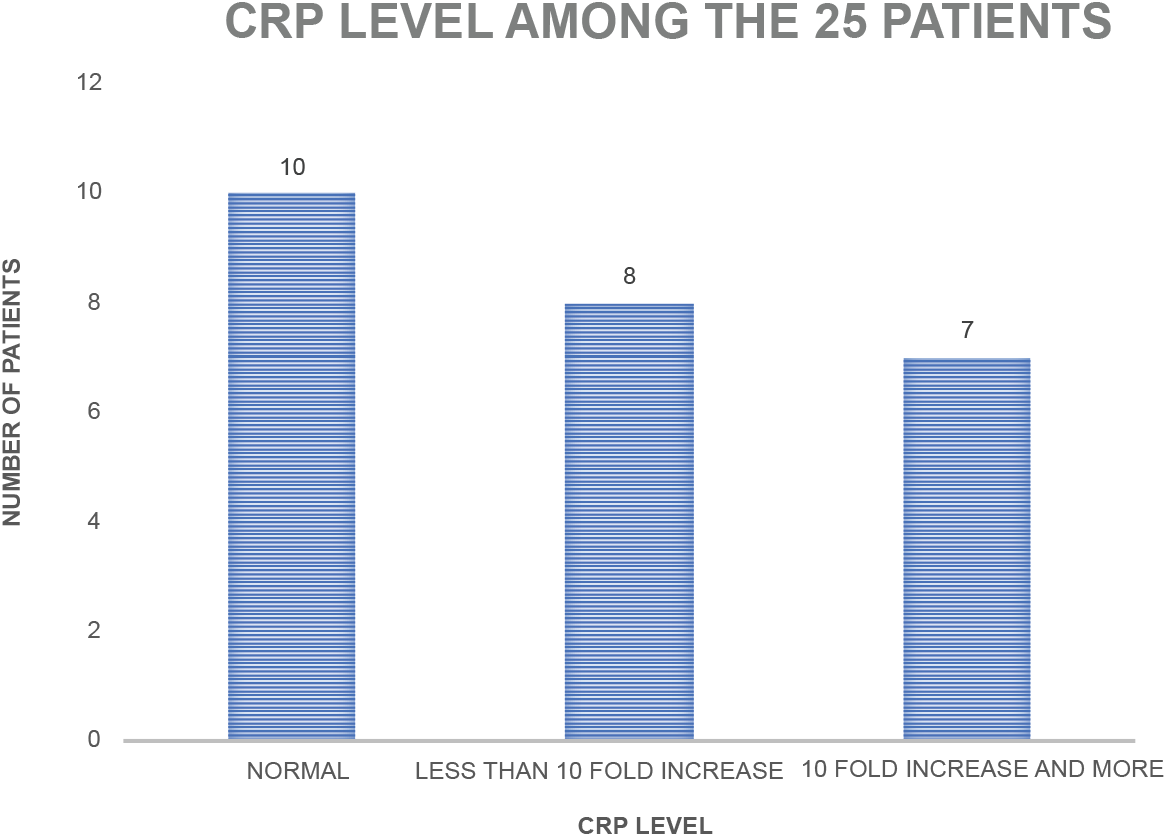

**FIGURE:5.**
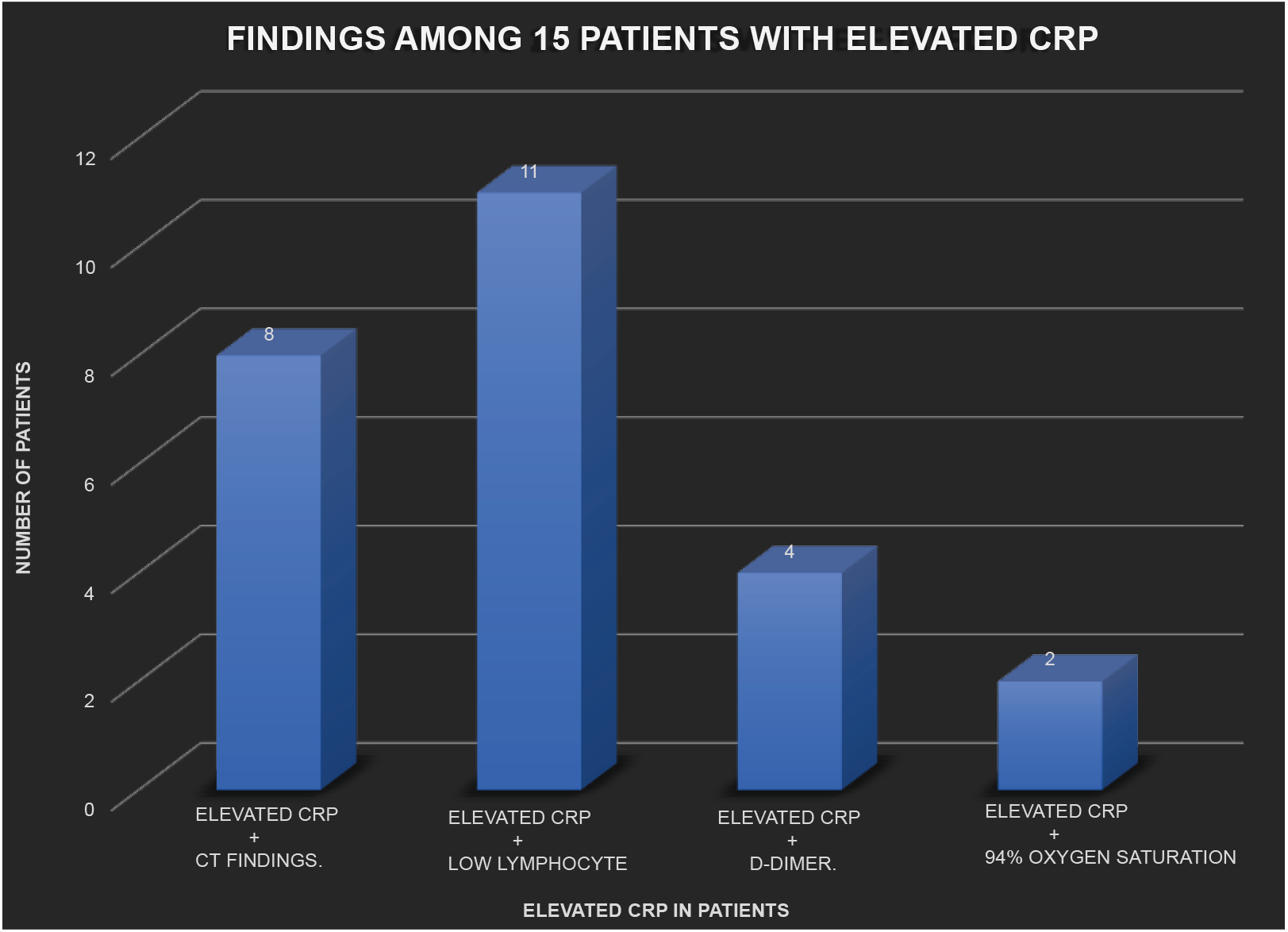

**FIGURE:6.**
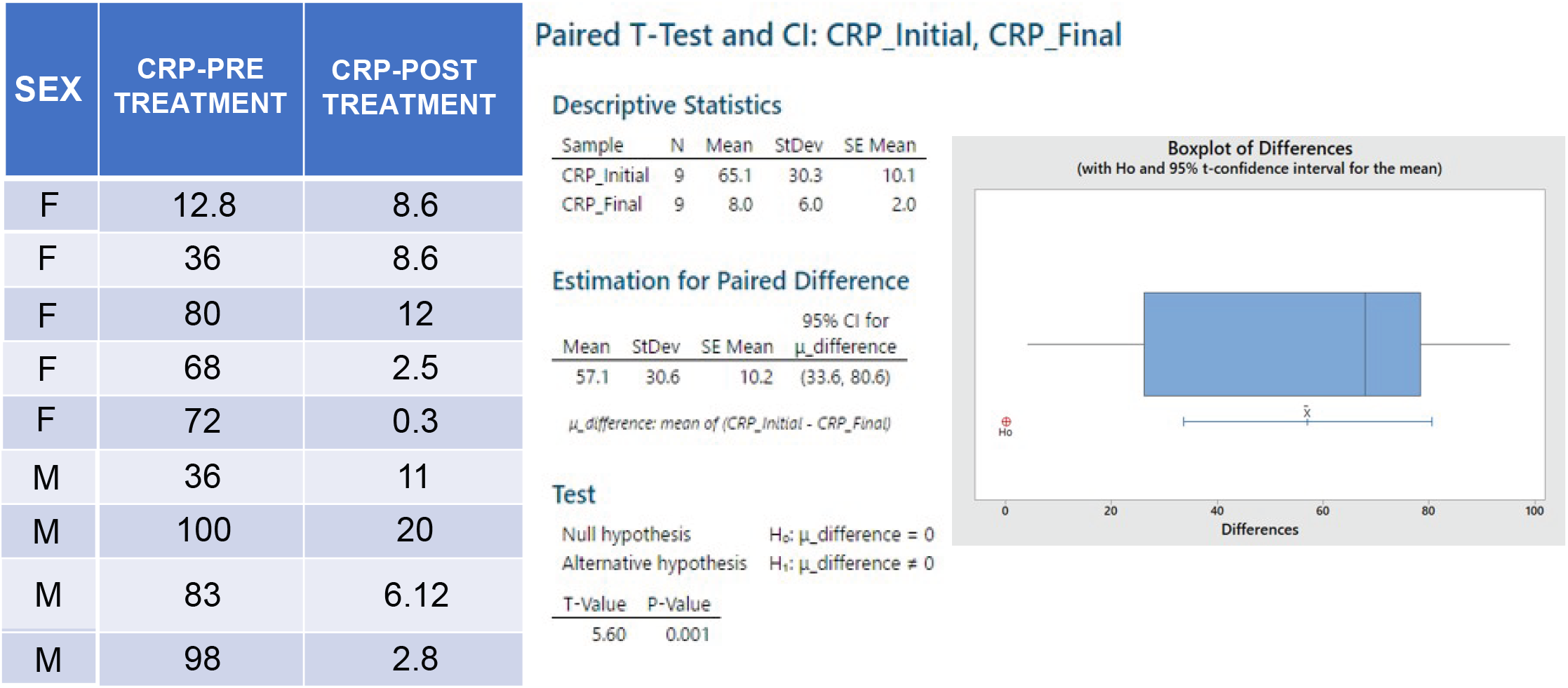

Among the 25 patients, 40% of the patients (10/25) had mild symptoms with normal CRP. 24% of the patients (6/25) had mild symptoms with elevated CRP. 36% of patients (9/25) had elevated CRP with persistent high fever.

Further analysis of CRP showed that 40% of the patients (10/25) had normal CRP and 60%of the patients (15/25) had an elevated CRP during the initial test. Out of this 60% of the patients (15/25), 32% of patients (8/25) had less than ten-fold increase in CRP level and 28% of patients (7/25) had ten-fold and more increase in CRP level.

Among the 24% patients (6/25) with mild symptoms and elevated CRP, 83.3% of the patients (5/6) had less than 10-fold increase in CRP and 16.7 % of the patients (1/6) had more than 10-fold increase in CRP. Among the 36% patients (9/25) with high grade fever and lower respiratory tract symptoms, 66.7% of the patients (6/9) had more than 10-fold increase in CRP.

Among the sub-group of 15 patients with elevated CRP, 73.3% patients (11/15) with elevated CRP also had low lymphocyte counts.,53.3% of the patients (8/15) had positive CT findings with CT severity score ranging from 8/25 to 15/25, (26.6%) of the patients (4/15) had elevated D-Dimer and 13.33% of the patients (2/15) had oxygen saturation 94% at the time of initial reporting.

During follow up of mildly symptomatic patients with elevated CRP, the CRP normalized with marked improvement of symptoms over the next three days of initiating treatment. In those 9 patients with high CRP and persistent high fever, there was a significant decrease in CRP in the immediate follow up test following early initiation of treatment. A paired sample t test using Minitab showed a p value of 0.001 indicating a significant difference in the CRP values post treatment. Only in 33.3% of the patients (3/9), the CRP values touched the baseline in the first follow up. In the rest of the patients, the CRP values remained low but did not touch the baseline even on the second follow up. In this group, all patients became afebrile within 24 hours of IV ceftriaxone and oral or IV steroid. But other symptoms like body pain, fatigue, loss of appetite and cough improved gradually over next one to three weeks.

All the 25 patients recovered over the next four weeks.

## DISCUSSION

The pathology of COVID 19 has an inflammatory and a thrombotic component^2,3,4^. COVID-19 is characterized by distinct changes in hematological, biochemical and radiological findings. Low lymphocyte count, raised levels of C reactive protein, lactate dehydrogenase and ferritin have been associated with more severe form of the disease.

We, therefore planned our early intervention based on CRP levels. CRP as a marker of inflammation was chosen as it has been shown as a predictor of disease severity in many studies and the test is readily available. Hospitalized patients who died from COVID-19 had about 10-fold increase in levels of CRP than those who recovered^5^. CRP in severe COVID-19 patients was reported to be significantly increased at the initial stage itself, even before CT findings^6^. CRP levels have been reported as a marker to predict the risk of COVID-19 aggravation in non-severe hospitalized adult patients^7^.

CT thorax was done for cases who had fever beyond third day and for patients who had persistent or productive cough.

Though WHO had initially not approved the use of steroids in COVID-19, the Recovery trial that evaluated potential treatments in hospitalized COVID-19 patients concluded that the use of dexamethasone resulted in lower 28-day mortality among those who were receiving either invasive mechanical ventilation or oxygen alone at randomization, but not among those receiving no respiratory support^8^. It has also been proposed that the timing and dose of steroids may play a role in preventing cytokine storm^9^.On the one hand, there are concerns that starting steroids at a very early stage of illness during viral replication may worsen the disease. On the other hand, starting steroids in patients already having hypoxia may not reverse the pathophysiology and clinical outcome in many cases. Large randomized trial data is also not available regarding the optimal timing for initiating steroids in symptomatic COVID-19 patients. Therefore, we started steroids at that stage of illness, when the inflammatory phase was waxing and the viral replication phase was waning in order to prevent complications arising from inflammation and cytokine storm.

Ceftriaxone is a prescribed drug under COVID-19 protocol. It is indicated to treat secondary community associated pneumonia^10^. It is reported to reduce neuronal hyperexcitability by reducing glutamate and crosses the blood brain barrier^11^. It is also reported to alter cytokine response^12^. We therefore used ceftriaxone for treating patients with persistent high-grade fever on the third and fourth day of illness.

The following protocol was followed for all the home quarantined patients in whom oxygen saturation was 94 percent and above.:

1. For patients with mild symptoms and normal CRP level, oral antibiotic Azithromycin 500mg od, Cetirizine 10mg at bedtime, Paracetamol and multi-vitamins were given. Mild cases were defined as those patients who presented with low grade fever of one day duration, mild body pain, nasal stuffiness and upper respiratory tract symptoms. CT scan was not done in these cases as it was not indicated. Steroid was not given in these patients as CRP levels were normal and there was no evidence of significant inflammation.
2. For patients with mild symptoms and less than ten-fold increase in CRP, low dose oral methyl prednisolone (8mg bd) was added. Low dose steroid was used because these patients were presumed to have mild inflammatory body response as evidenced by the CRP levels. CT scan was not done as there was no indication.
3. For patients with high grade fever persisting even on third or fourth day after onset of symptoms or 10-fold or more increase in CRP, intravenous ceftriaxone with either oral methyl prednisolone or intravenous dexamethasone was given
  a. The dose of Injection ceftriaxone was 1gm bd.
  b. The dose of oral methylprednisolone was 16 mg bd or 32mg per day. Maximum dose was given for 5 days and tapered over next 4 to 5 days
  c. The dose of IV dexamethasone was 6 mg bd. This was given to patients who could not tolerate oral steroids.
  d. For most patients in this group, CT scan was indicated.
4. Rivaroxaban 5mg, or 10mg was given for minimum 4 weeks depending on D dimer levels.
5. After the first online consultation on the third or fourth day after onset of symptoms, all the patients were followed-up through online consultation for four weeks.

With the above treatment model, we observed that

1. All the mildly symptomatic patients with normal CRP who received oral antibiotics and no steroid recovered as anticipated. Their oxygen saturation remained normal throughout the course of the illness and during the follow up.
2. Patients with mild symptoms and less than 10-fold increase in CRP, who received low dose oral steroid and oral antibiotic improved symptomatically. There were no abnormalities in oxygen saturation throughout the course of illness and during the follow up.
3. To our surprise, we noted that patients who appeared toxic initially with high grade fever or ten-fold or more increase in CRP showed marked improvement of their febrile status within 24 hours. However, body pain, fatigue and loss of appetite improved but persisted over next seven to ten days. Dry cough persisted for three to four weeks. None of the patients with toxic appearance, high fever and markedly elevated CRP who were treated on their third or fourth day of illness worsened.

Two patients with high CRP had reported with a low oxygen saturation of 94% on their first consultation. After initiation of treatment, their saturation improved overnight to normal limits. During the course of treatment of patients with high fever and high CRP, two patients had a drop in oxygen saturation to 94% on the third day after the start of the treatment which lasted for a few hours and returned to normal within a few hours. The oxygen saturation level of all the remaining patients remained normal. None of the cases developed thrombotic events during the treatment.

This group of patients are those who would have probably deteriorated during home quarantine while waiting for the danger signs and symptoms to appear and would have contributed to late presentation to hospital and increased mortality.

If these patients had not been monitored at home in the initial stages with CRP and D dimer values, some of them might have died of hypoxia due to pulmonary complications or thrombotic events.

Till date, no data is available on the morbidity and mortality rate of home quarantined patients. During the second wave of COVID-19 in India, many COVID hospitals witnessed home quarantined patients either being brought dead or brought in a very advanced stage of illness. A large randomized study on home quarantined patients with elevated inflammatory markers and persistent fever will through light on the mortality and morbidity benefits of early intervention.

Based on our experience of treating a small group of COVID patients under home quarantine we also realized that

1. Patients who were initially classified as mild and advised to be on home quarantine on their first visit to the COVID hospital and those patients who preferred to stay at home thinking that they had only mild disease, did not remain mild towards the end of the first week.
2. Many of our patients did not know the severity of their symptoms and were unaware when to contact the hospital. This is because except for oxygen saturation monitoring, all other danger signs and symptoms were subjective
3. Many patients were either reluctant or delayed getting admitted in the hospital because of the overcrowding in COVID hospitals. This led to worsening of their illness to a point of no recovery.
4. Few were worried about the stigma of being diagnosed with the viral illness. Fearing criticism, they did not report to hospital till they became very sick.
5. Many patients who are in home isolation are unable to reach out to their relatives for timely help because of social constraints.
6. Early intervention, by improving clinical status and preventing fall in oxygen saturation gave a sense of confidence and positive thinking to the patients
7. Only those patients who were lucky to have access to a family physician like the present group of patients had the opportunity to survive.

Therefore, this model of home quarantine approach under the supervision of a trained medical professional can save lives. This model can be easily replicated at the community level and even in rural settings with limited resources.

1. A limited number of patients in a locality can be provided online access to a medical professional who can monitor the patients during the home quarantine.
2. Mobile units can provide specific laboratory tests for regular monitoring.
3. The local community halls and primary health centers can also be used for dispensing medication as per medical advice.
4. This model could be useful in micro-managing COVID positive patients at their homes and thereby avoid crowding of the COVID centers.
5. Home quarantine and intake of low dose steroids at home can also avoid incidence of nosocomial infections like mucormycosis.

## CONCLUSION

The existing guidelines initiate monitoring of inflammatory markers and starting of interventions only after the onset of hypoxia in hospital settings.

We propose (Fig:1) an early objective assessment of inflammatory markers like CRP prior to the onset of hypoxia during home quarantine and early, need-based intervention under medical guidance to avoid complications and hospitalization.

## Data Availability

Data analyzed during the study are included in the article.

## Acknowledgement

We acknowledge Mr. Arjun Pratapchandran for his invaluable support in analyzing and formatting the study data.

## Data Availability

Data analyzed during the study are included in the article.

## REFERENCES

1. Revised guidelines for Home Isolation of mild /asymptomatic COVID-19 cases dated 28 April 2021. https://www.mohfw.gov.in/pdf/Guidelinesforhomequarantine.pdf.

2. Sriram K; Insel P.A; Inflammation and thrombosis in COVID-19 pathophysiology: proteinase-activated and purinergic receptors as drivers and candidate therapeutic targets; 2021, Physiol Rev 101: 545–567

3. Giovanni Ponti, Monia Maccaferri, Cristel Ruini, Aldo Tomasi & Tomris Ozben (2020) Biomarkers associated with COVID-19 disease progression, Critical Reviews in Clinical Laboratory Sciences, 57:6, 389–399, DOI: 10.1080/10408363.2020.1770685

4. Tassiopoulos AK, Mofakham S, Rubano JA, Labropoulos N, Bannazadeh M, Drakos P, Volteas P, Cleri NA, Alkadaa LN, Asencio AA, Oganov A, Hou W, Rutigliano DN, Singer AJ, Vosswinkel J, Talamini M, Mikell CB and Kaushansky K (2021) D-Dimer-Driven Anticoagulation Reduces Mortality in Intubated COVID-19 Patients: A Cohort Study with a Propensity-Matched Analysis. Front. Med. 8:631335. doi: 10.3389/fmed.2021.631335

5. Ali N. (2020). Elevated level of C-reactive protein may be an early marker to predict risk for severity of COVID-19. Journal of medical virology,92(11), 2409–2411.

6. Tan, C., Huang, Y., Shi, F., Tan, K., Ma, Q., Chen, Y., Jiang, X., & Li, X. (2020). C-reactive protein correlates with computed tomographic findings and predicts severe COVID-19 early. Journal of medical virology,92(7), 856–862.

7. Wang G, Wu C, Zhang Q, Wu F, Yu B, Lv J, Li Y, Li T, Zhang S, Wu C, Wu G, Zhong Y. C-Reactive Protein Level May Predict the Risk of COVID-19 Aggravation. Open Forum Infect Dis. 2020 Apr 29;7(5): ofaa153.

8. The RECOVERY Collaborative Group. Dexamethasone in Hospitalized Patients with COVID-19; N Engl J Med 2021; 384; 8:693–704.

9. Ur A, Verma K. Cytokine Storm in COVID19: A Neural Hypothesis. ACS Chem Neurosci. 2020 Jul 1 ;11(13) :1868–1870. doi: 10.1021/acschemneuro.0c00346.

10. Nestler MJ, et al. (2020). Impact of COVID-19 on pneumonia-focused antibiotic use at an academic medical center. Infection Control & Hospital Epidemiology, https://doi.org/10.1017/ice.2020.362

11. Yimer EM, Hishe HZ and Tuem KB (2019) Repurposing of the β-Lactam Antibiotic Ceftriaxone for Neurological Disorders: A Review. Front. Neurosci. 13: 236.doi: 10.3389/fnins.2019.00236.

12. Anuforom O, Wallace GR, Buckner MMC and Piddock LJV (2016) Ciprofloxacin and ceftriaxone alter cytokine responses, but not Toll-like receptors, to Salmonella infection in vitro J Antimicrob Chemother, Jul;71(7):1826–33.

